# Two-Month Follow-up of Persons with SARS-CoV-2 Infection—Zambia, September 2020

**DOI:** 10.1101/2021.06.15.21258964

**Authors:** James E. Zulu, D. Banda, J.Z. Hines, M. Luchembe, S. Sivile, M. Simwinga, D. Kampamba, K. Zyambo, R. Chirwa, L. Chirwa, W. Malambo, D.T. Barradas, N. Sinyange, S. Agolory, L.B. Mulenga, S. Fwoloshi

## Abstract

**Background:** COVID-19 is often characterized by an acute upper respiratory tract infection. However, information on longer-term clinical sequelae following acute COVID-19 is emerging. We followed a group of persons with COVID-19 in Zambia at two months to assess persistent symptoms.

**Methods:** In September 2020, we re-contacted participants from SARS-CoV-2 prevalence studies conducted in Zambia in July 2020 whose PCR tests were positive. Participants with valid contact information were interviewed using a structured questionnaire that captured demographics, pre-existing conditions, and types and duration of symptoms. We describe the frequency and duration of reported symptoms and used chi-square tests to explore variability of symptoms by age group, gender, and underlying conditions.

**Results:** Of 302 participants, 155 (51%) reported one or more acute COVID-19-related symptoms in July 2020. Cough (50%), rhinorrhoea (36%) and headache (34%) were the most frequently reported symptoms proximal to diagnosis. The median symptom duration was 7 days (IQR: 3-9 days). At a median follow up of 54 days (IQR: 46-59 day), 27 (17%) symptomatic participants had not yet returned to their pre-COVID-19 health status. These participants most commonly reported cough (37%), headache (26%) and chest pain (22%). Age, sex, and pre-existing health conditions were not associated with persistent symptoms.

**Conclusion:** A notable percentage of persons with SARS-CoV-2 infection in July still had symptoms nearly two months after their diagnosis. Zambia is implementing ‘post-acute COVID-19 clinics’ to care for patients with prolonged symptoms of COVID-19, to address their needs and better understand how the disease will impact the population over time.

## Introduction

Since detecting the first case of Coronavirus disease 2019 (COVID-19) in March 2020, Zambia has experienced substantial transmission of SARS-CoV-2, the virus that causes COVID-19. From March 18, 2020 to March 30, 2021, Zambia had reported 88,012 cases to the country’s COVID-19 surveillance system. Modelling projections estimate the true number of infections to be at least 10 times greater than this figure (1–3).

As a new disease, the full clinical spectrum and sequelae of COVID-19 is still emerging. Describing the natural history of SARS-CoV-2 infection is important to ensure essential health services and appropriate budget allocation (4). Globally, estimates of asymptomatic SARS-CoV-2 infections range from 20% to 70%(5). When symptoms occur, they typically appear between 2 to 14 days following exposure; fever, dry cough, and shortness of breath are common. Acute symptoms may persist for up to two additional weeks(6,7). Ongoing symptomatic COVID-19 involves signs and symptoms of COVID-19 that occur from 4 to 12 weeks, while Post-COVID-19 syndrome (“long COVID”) is described as signs and symptoms that develop during or after an infection consistent with COVID-19 that are not explained by an alternative diagnosis and continue for more than 12 weeks (8). Symptom severity can vary by age and pre-existing health conditions (9).

Long-term health effects of COVID-19 have been reported(10,11). Patients hospitalized with COVID-19 often had persistent symptoms, abnormal patterns on chest imaging, impaired lung function, and poor quality of life for several weeks to months following infection(12–14). In one study from Italy, more than a third of hospitalized patients with COVID-19 experienced more than one persistent symptom (12). The most frequently reported long-term symptoms included fatigue (range: 15-87%), dyspnea (range: 10-71%), chest-discomfort (range: 12-44%) and cough (range: 17-26%) (12,15–20). Although less common, persons with mild and moderate COVID-19 have also reported long-term health effects. Of 669 non-hospitalized patients in a study in Geneva, Switzerland, 32% reported fatigue, dyspnea and loss of taste or smell 30-45 days following symptom onset (19). Information on long-term sequelae following SARS-CoV-2 infections in Africa is limited. In this project, we interviewed persons previously diagnosed with laboratory confirmed SARS-CoV-2 infection to describe their long-term experience with COVID-19 related symptoms. We anticipate that findings from this project can be used to inform resource requirements for additional care and support as the COVID-19 pandemic evolves.

## Methods

We contacted participants who tested positive for SARS-CoV-2 during prevalence studies conducted previously in Zambia. These previous studies were conducted in Kabwe, Livingstone, Lusaka, Nakonde, Ndola, and Solwezi Districts in July 2020. These districts have a combined population of 4,290,107 persons, one-quarter of Zambia’s population(21). The districts were purposefully selected based on high rates of SARS-CoV-2 infections reported in official Ministry of Health statistics (80% of laboratory-confirmed cases in Zambia were reported from these six districts from March to June 2020) and being highly populated areas, transit corridors and/or points-of-entry (3). Participants were recruited from the general population (N=4,258), outpatient department attendees (N=1,952), and health care workers (N=660) regardless of symptoms. Each participant was tested for SARS-CoV-2 infection by real-time polymerase chain reaction (PCR) using nasopharyngeal specimens. Patients who were hospitalized were not included in the prevalence studies. This follow-on study was approved by the Zambia National Health Research Authority and the University of Zambia Biomedical Research Ethics Committee. The activity (initial prevalence study and this study) was reviewed by CDC and was conducted consistent with applicable federal law and CDC policy.^*^

For the current project, we identified previous prevalence study participants with valid contact information. Eleven project staff members contacted these participants in September 2020 (approximately two months following the end of the prevalence survey in July 2020). If the participant provided consent, project staff conducted a 15-20 minute interview using a structured questionnaire. The questionnaire captured demographic information, pre-existing comorbid conditions, types and duration of COVID-19 symptoms, when participants returned to their previous health status, and whether participants visited a health facility due to their symptoms. Because participants were only assessed at one time point after initial diagnosis, the symptom duration for those with persistent symptoms could not be assessed. Project staff attempted to reach each participant by phone up to five times, after which staff visited the participants’ home. Responses were recorded on tablets and entered into a REDCap database maintained by the Zambia Ministry of Health. Data were then imported into STATA version 14.2 for analysis (22).

We defined persistent symptoms as participants having continual symptoms since diagnosis with SARS-CoV-2 infection until time of interview. Persons with symptoms duration of less than 4 weeks were not considered persistent. The term “post-COVID syndrome” was deliberately not used to describe these persons because our assessment occurred before the 12-week timeline as recommended by some authorities (8). We described the frequency and duration of reported symptoms and used equality of median test and chi square test to explore variability of symptoms at onset, persistent symptoms, and symptom duration by age group, gender, and underlying conditions. We conducted bivariate logistic regression to calculate the odds of having persistent symptoms among symptomatic patients by age, sex, comorbidities, number of symptoms at onset, and specific symptoms at onset. An alpha level of 0.05 was used to assess statistical significance.

## Results

Overall, 525 (7.6%) participants had positive SARS-CoV-2 PCR tests in the prevalence studies from July 2020. Of these, 431 (82%) participants had valid contact information and 327 (62%) participants were reached by phone or in-person for this study. The final sample included 302 (58%) participants who agreed to be interviewed. The median duration between positive COVID-19 test and follow up was 54 days [interquartile range (IQR): 46-59 days]. Mean participant age was 32 years (range: 1-85 years) and 42% were males (Table 1). Eighteen percent reported any comorbidity, with HIV (8%) and hypertension (5%) being the two most common conditions reported.

**Table 1.** Demographic characteristics of persons with positive SARS-CoV-2 PCR test by initial and persistent symptom status—Zambia, July 2020

|  |  | Overall<br>N=302 | Initial<br>Symptoms* N=155 | Asymptomatic<br>N=147 |  | Persistent<br>symptoms<br>N=27<sup>†</sup> | Resolved<br>symptoms<br>N=125<sup>†</sup> |  |
| --- | --- | --- | --- | --- | --- | --- | --- | --- |
|  |  | N (%) | N (%) | N (%) | P-Value | N (%) | N (%) | P-Value |
| Symptomatic |  | 155 (51.3) |  |  |  |  |  |  |
| Sex | Male | 128 (42.4) | 64 (41.3) | 64 (43.5) | 0.693<sup>¶</sup> | 10 (37.0) | 53 (42.4) | 0.818<sup>¶</sup> |
|  | Female | 174 (57.6) | 91 (58.7) | 83 (56.5) |  | 17 (63.0) | 72 (57.6) |  |
| Age Group | 0-19 | 63 (21.0) | 35 (22.9) | 28 (19.0) | 0.718<sup>¶</sup> | 6 (22.2) | 27 (22.0) | 0.692** |
|  | 20-49 | 195 (65.0) | 97 (63.4) | 98 (66.7) |  | 18 (66.7) | 79 (64.2) |  |
|  | 50+ | 42 (14.0) | 21 (13.7) | 21 (14.3) |  | 3 (11.1) | 17 (13.8) |  |
| Comorbidity<sup>‡</sup> |  | 54 (17.9) | 21 (13.6) | 33 (22.4) | 0.044<sup>¶</sup> | 3 (11.1) | 18 (14.4) | 0.672** |
| HIV |  | 23 (8.1) | 9 (6.0) | 14 (10.3) | 0.188<sup>¶</sup> | 2 (7.4) | 7 (5.9) | 0.716** |
| Hypertension |  | 16 (5.3) | 6 (3.9) | 10 (6.8) | 0.309<sup>¶</sup> | 0 (0.0) | 6 (4.8) | 0.407** |

\* Includes those that were experienced at time of diagnosis as well as those that developed after diagnosis
<sup>†</sup> Three persons (one male, two females: 2 below the age of 20 years and one above 50 years), who indicated 'don't know' if they had persistent symptoms were excluded
<sup>‡</sup> Comorbid conditions included: Hypertension, HIV, Diabetes Mellitus, Asthma, Chronic Obstructive Pulmonary Disease and Cancer
<sup>¶</sup> Pearson Chi-square test
\*\* Fisher's exact test
PCR: Polymerase Chain reaction

Of 302 study participants, 155 (51%) developed any symptoms (Table 1). Most (69%) were symptomatic at the time of testing while the remainder (31%) developed symptoms after testing. The median duration of symptoms for those without persistent symptoms was 7 days (IQR: 3-9 days). There were no differences in duration of symptoms by age or sex but, paradoxically, those with comorbid conditions reported a shorter median symptom duration (2 days [IQR: 2-4 days]) compared to those without comorbid conditions (7 days [IQR: 5-7 days]) (p=0.04).

The most common symptoms reported around the time of diagnosis were cough (50%), rhinorrhoea (36%), headache (34%), fatigue (29%) and fever (20%) (Table 2). Cough (22%) was the most commonly reported first symptom. Forty-four (28%) symptomatic participants sought care for their symptoms in a health facility. Participants with comorbid conditions were more likely to be symptomatic (p=0.044), whereas age and sex were not associated with being symptomatic (Table 1).

**Table 2.**
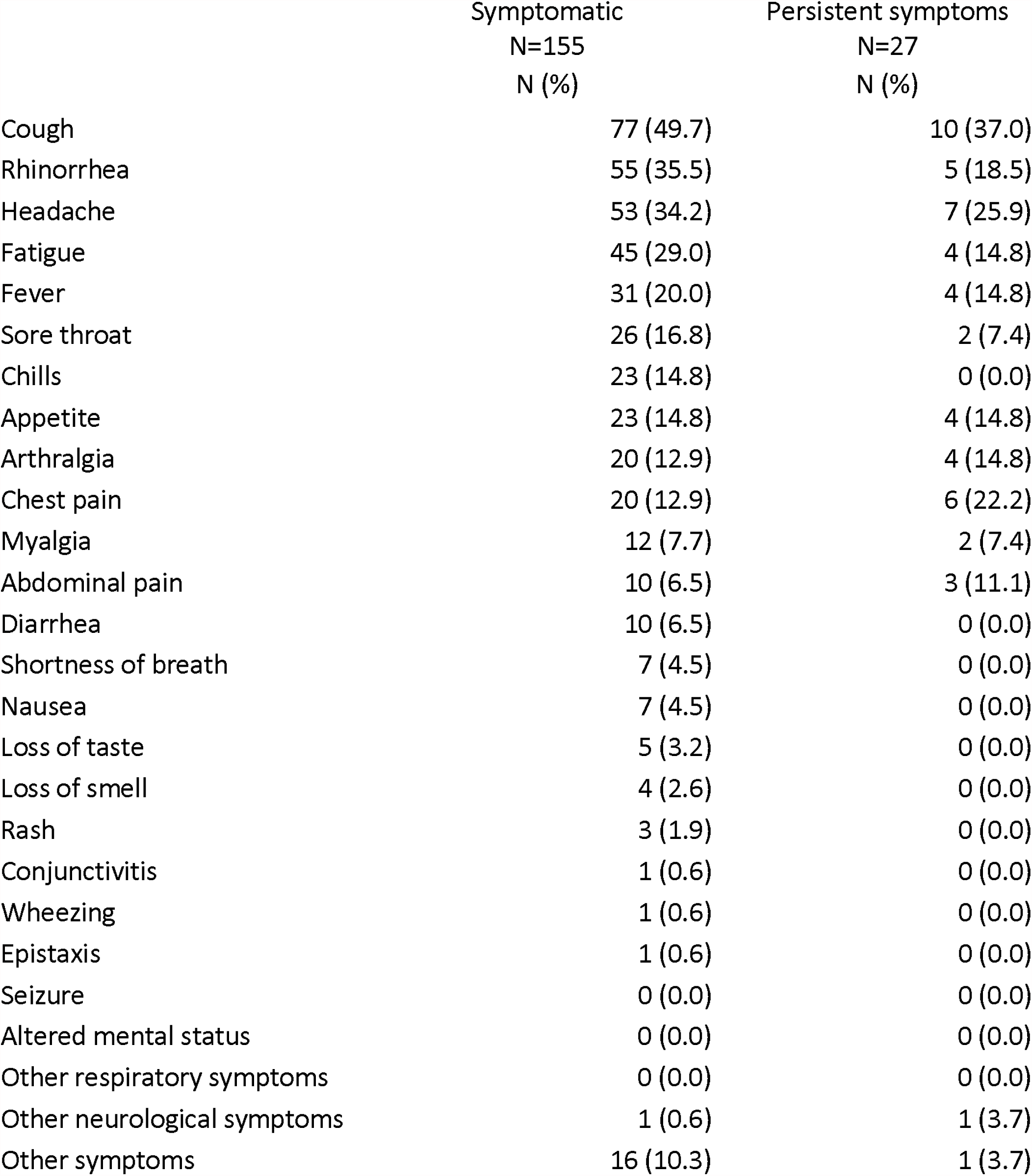
Long term symptomology of persons with positive SARS-CoV-2 PCR test—Zambia, July 2020

Overall, 27 (17%) symptomatic participants had not returned to their usual state of health at time of interview for this study. The median age of participants with persistent symptoms was 33 years [IQR: 25-42] and 17 (63%) were females. Two (7%) participants with persistent symptoms reported being infected with HIV, one reported (4%) having diabetes mellitus, while the rest did not report any comorbidities. The most common ongoing symptoms were cough (37%), headache (26%), chest pain (22%), and rhinorrhoea (19%) (Table 2). Six (22%) were 19 years of age or younger, including two males and four females; they mainly complained of cough, fever, chills, arthralgia, headache, and rhinorrhoea among other symptoms. Age, sex, and having any comorbidity were not associated with having persistent symptoms (p≥0.05 for all) (Table 1). However, on bivariate logistic regression analysis, those with five or more symptoms at onset had nearly three-times the odds of reporting persistent symptoms compared other symptomatic participants (p=0.03) (Table 3). Participants with loss of appetite at symptoms onset also had higher odds of reporting persistent symptoms (p=0.01).

**Table 3.** Odds ratios for factors associated with experiencing persistent symptoms among persons with positive SARS-CoV-2 PCR test—Zambia, September 2020

| Characteristic | OR | p-value | 95% CI |
| --- | --- | --- | --- |
| **Sex** |  |  |  |
| Male | 1.27 | 0.567 | 0.56-2.89 |
| Female | Ref. |  |  |
| **Age** |  |  |  |
| 0-19 | Ref. |  |  |
| 20-49 | 0.77 | 0.584 | 0.30-1.96 |
| ≥50 | 0.79 | 0.737 | 0.21-3.05 |
| **Past medical history** |  |  |  |
| HIV | 1.14 | 0.872 | 0.23-5.80 |
| Diabetes | 4.28 | 0.309 | 0.26-70.4 |
| Hypertension | 1.00 | - | - |
| comorbidity | 0.88 | 0.814 | 0.30-2.55 |
| **Number of Symptoms at onset** |  |  |  |
| 1-2 | Ref. |  |  |
| 3-4 | 1.09 | 0.863 | 0.40-2.98 |
| ≥5 | 2.87 | 0.032 | 1.09-7.56 |
| **Symptoms at onset** |  |  |  |
| Cough | 1.42 | 0.395 | 0.63-3.16 |
| Rhinorrhea | 0.39 | 0.054 | 0.15-1.02 |
| Headache | 1.14 | 0.751 | 0.50-2.62 |
| Sorethroat | 1.31 | 0.599 | 0.48-3.61 |
| Chills | 2.07 | 0.151 | 0.77-5.61 |
| Loss of appetite | 3.40 | 0.012 | 1.30-8.86 |
| Arthralgia | 2.62 | 0.065 | 0.94-7.29 |
| Fever | 1.00 | 1.000 | 0.37-2.71 |
| Loss of taste | 2.90 | 0.255 | 0.46-18.2 |
| **Sought Care** | **0.57** | **0.204** | **0.23-1.36** |

OR- Odds ratio, AOR- adjusted Odds ratio, CI- confidence interval.

## Discussion

Only half of persons with SARS-CoV-2 infection were symptomatic, and among those with symptoms, a notable portion were still experiencing symptoms nearly two months after testing positive in this non-hospitalized cohort. The World Health Organization estimates recovery time for SARS-CoV-2 infection is two weeks for mild infections and three to six weeks for severe infections (23), recognizing the recovery course can be variable (24), and that post-COVID-19 syndrome is an emerging public health problem (25). This study adds to the growing evidence that COVID-19 symptoms can persist beyond the acute infection period, even among persons with less severe infection, and, to our knowledge, is the first from Africa to report such findings. Furthermore, in this study chronic cough was the most common persistent symptom, which even among persons with a history of COVID-19 warrants further clinical investigation, particularly in high tuberculosis burden regions like sub-Saharan Africa.

Several studies from the United States (U.S.) and Europe have demonstrated that symptoms can persist beyond two weeks among outpatients with COVID-19 (10,11). In a U.S. study of 150 patients with non-critical COVID-19 followed up at 60 days after symptom onset, two-thirds were symptomatic [10]. In a survey of 292 patients diagnosed with COVID-19 in an outpatient setting in the U.S., only 65% reported a return to baseline health by 14 to 21 days after diagnosis (11). Those who did return to baseline health did so a median of seven days after diagnosis, which is comparable to the current study. The persistent symptom profile included similar symptoms as reported in this study (i.e., cough and fatigue) as well as differences (i.e., anosmia, ageusia, dyspnoea, and asthenia) (10,11). Additionally, differences in the symptomatic rate at testing as well as a proportion of persons with ongoing symptoms were reported. Furthermore, persistent symptoms are recognized as a problem for paediatric patients (26), as was also observed in this study. Lastly, data from 4,182 persons with COVID-19 in the United Kingdom (UK), United States and Sweden, using self-reported symptoms into a COVID-19 symptom tracking app found that experiencing more than five symptoms during the first week of illness was associated with long COVID-19 (27), which is similar to our study.

Hospitalized patients appear to report higher prevalence of persistent symptoms than outpatients. In a survey of 350 hospitalized patients with COVID-19 in the U.S., only 39% reported a return to baseline health by 14 to 21 days after diagnosis (28). Similarly, a study of 143 patients who had been hospitalized for COVID-19 in Italy, found that only 13% were symptom free after a mean of 60 days following disease onset (12). Hospitalized patients with more severe illness have reported experiencing symptoms for several months (29). A cohort study of hospitalized persons with COVID-19 in Wuhan, China concluded that at six months after symptom onset, patients were still reporting fatigue, muscle weakness, and sleep difficulties as the residual symptoms(29).

Systematic evaluations of long-term physiological effects of COVID-19 are lacking, but emerging data (10,15,30) and evidence from other coronaviruses [28] suggest the potential for ongoing respiratory impairment. Moreover, cardiac and pulmonary diagnostic studies suggest the potential for long-term sequelae after COVID-19, even among patients managed in the outpatient setting (15,30).

In this study, persons with symptomatic infections commonly reported non-specific acute COVID-19 symptoms, while fever and shortness-of-breath—two of the three classic symptoms of COVID-19 (31)—were rarely reported. This finding was consistent with a UK Biobank SARS-CoV-2 serology study which found that 40% of the seropositive participants did not have one of the three classic COVID-19 symptoms (32). These findings point to the need for clinicians to maintain a high index of suspicion and low testing threshold for COVID-19 when high levels of SARS-CoV-2 transmission are occurring in a community.

This study was subject to several limitations. Firstly, this was a convenience sample of the general population, outpatients, and health care workers who tested positive and therefore may not be representative of all persons with COVID-19 in Zambia. Furthermore, the response rate was low and persons with persistent symptoms might have had greater (or less) participation than other participants. The data were collected approximately two months following the participants’ positive SARS-CoV-2 test result, hence responses were subject to recall bias. The symptoms and return to usual health were self-reported and may not accurately represent a comparable picture of COVID-19 persistent symptoms. It is also possible that self-reported symptoms were caused by other conditions. Pre-existing comorbid conditions may have been mis-reported.

This study adds to a growing body of literature on long term clinical course of COVID-19 in non-hospitalized persons. It is becoming clear that COVID-19 symptoms can persist for weeks to months. Given the extent of the outbreak in Africa, the COVID-19 pandemic could create health problems for years to come. Governments need to proactively establish care models for post-acute COVID-19 care in Africa. In high tuberculosis burden countries like Zambia, persons with chronic cough should be assessed for tuberculosis infection. Zambia is implementing ‘post-acute COVID-19 clinics’ to care for patients with prolonged symptoms of COVID-19 to better address their health needs, to describe the natural history of COVID-19 in Zambia, and better understand how the disease will impact the Zambian population over time. Information gleaned from these clinics will help inform health services planning in the country.

## Data Availability

All data referred to in this manuscript is available for viewing and verification

## Conflicts of Interest

The authors declare no conflicts of interest with respect to this study

## Authorship Disclaimer

The findings and conclusions in this report are those of the authors and do not necessarily represent the official position of the Centers for Disease Control and Prevention of the Zambia Ministry of Health.

## Funding

This work was supported by the CDC Emergency Response to the COVID-19 pandemic and the President’s Emergency Plan for AIDS Relief (PEPFAR) through the Centers for Disease Control and Prevention (CDC).

## Footnotes

* See e.g., 45 C.F.R. part 46, 21 C.F.R. part 56; 42 U.S.C. §241(d); 5 U.S.C. §552a; 44 U.S.C. §3501 et seq.

## Notes

### Competing Interest Statement

The authors have declared no competing interest.

### Funding Statement

This study was funded by the US Centres for Disease Control and Prevention

### Author Declarations

The University of Zambia Biomedical Research Ethics Committee Ridgeway Campus, P.O. Box 50110 Lusaka, Zambia

## References

1. University of Basel. COVID-19 Scenarios. 2021; Based on inputs by Adam Wolkon at CDC-Zambia. Available upon request.

2. Pearson CAB, van Schalkwyk C, Foss AM, O’Reilly KM, Pulliam JRC. Projected early spread of COVID-19 in Africa through 1 June 2020. Eurosurveillance. 2020;25(18):1–6.

3. Mulenga LB, Hines JZ, Fwoloshi S, Chirwa L, Siwingwa M, Yingst S, et al. Prevalence of SARS-CoV-2 in six districts in Zambia in July, 2020: a cross-sectional cluster sample survey. Lancet Glob Heal. 2021;(21):1–9.

4. Yelin D, Wirtheim E, Vetter P, Kalil AC, Bruchfeld J, Runold M, et al. Long-term consequences of COVID-19: research needs. Vol. 20, The Lancet Infectious Diseases. 2020.

5. Yanes-Lane M, Winters N, Fregonese F, Bastos M, Perlman-Arrow S, Campbell JR, et al. Proportion of asymptomatic infection among COVID-19 positive persons and their transmission potential: A systematic review and meta-analysis. PLoS One. 2020 Nov;15(11).

6. Wiersinga WJ, Rhodes A, Cheng AC, Peacock SJ, Prescott HC. Pathophysiology, Transmission, Diagnosis, and Treatment of Coronavirus Disease 2019 (COVID-19): A Review. Vol. 324, JAMA - Journal of the American Medical Association. 2020.

7. Cevik M, Kuppalli K, Kindrachuk J, Peiris M. Virology, transmission, and pathogenesis of SARS-CoV-2. BMJ. 2020;371.

8. NICE. COVID-19 rapid guideline: managing the long-term effects of COVID-19 NICE guideline [NG188]. Natl Inst Heal Care Excell. 2020;(December 2020).

9. Centers for Disease Control and Prevention. Post-COVID Conditions [Internet]. 2021. Available from: https://www.cdc.gov/coronavirus/2019-ncov/long-term-effects.html

10. Carvalho-Schneider C, Laurent E, Lemaignen A, Beaufils E, Bourbao-Tournois C, Laribi S, et al. Follow-up of adults with noncritical COVID-19 two months after symptom onset. Clin Microbiol Infect [Internet]. 2020; Available from: https://doi.org/10.1016/j.cmi.2020.09.052

11. Tenforde MW, Kim SS, Lindsell CJ, Billig Rose E, Shapiro NI, Files DC, et al. Symptom Duration and Risk Factors for Delayed Return to Usual Health Among Outpatients with COVID-19 in a Multistate Health Care Systems Network — United States, March–June 2020. MMWR Morb Mortal Wkly Rep. 2020 Jul;69(30).

12. Carfì A, Bernabei R, Landi F. Persistent Symptoms in Patients After Acute COVID-19. JAMA. 2020 Aug;324(6).

13. Huang Y, Tan C, Wu J, Chen M, Wang Z, Luo L, et al. Impact of coronavirus disease 2019 on pulmonary function in early convalescence phase. Respir Res. 2020 Dec;21(1).

14. Liu C, Ye L, Xia R, Zheng X, Yuan C, Wang Z, et al. Chest Computed Tomography and Clinical Follow-Up of Discharged Patients with COVID-19 in Wenzhou City, Zhejiang, China. Ann Am Thorac Soc. 2020 Oct;17(10).

15. Xiong Q, Xu M, Li J, Liu Y, Zhang J, Xu Y, et al. Clinical sequelae of COVID-19 survivors in Wuhan, China: a single-centre longitudinal study. Clin Microbiol Infect. 2021 Jan;27(1).

16. Goërtz YMJ, Van Herck M, Delbressine JM, Vaes AW, Meys R, Machado FVC, et al. Persistent symptoms 3 months after a SARS-CoV-2 infection: the post-COVID-19 syndromeã ERJ Open Res. 2020 Oct;6(4).

17. Halpin SJ, McIvor C, Whyatt G, Adams A, Harvey O, McLean L, et al. Postdischarge symptoms and rehabilitation needs in survivors of COVID-19 infection: A cross-sectional evaluation. J Med Virol. 2021 Feb;93(2).

18. Wong AW, Shah AS, Johnston JC, Carlsten C, Ryerson CJ. Patient-reported outcome measures after COVID-19: a prospective cohort study. Eur Respir J. 2020 Nov;56(5).

19. Nehme M, Braillard O, Alcoba G, Aebischer Perone S, Courvoisier D, Chappuis F, et al. COVID-19 Symptoms: Longitudinal Evolution and Persistence in Outpatient Settings. Ann Intern Med. 2020 Dec;

20. Zhao Y, Shang Y, Song W, Li Q, Xie H, Xu Q, et al. Follow-up study of the pulmonary function and related physiological characteristics of COVID-19 survivors three months after recovery. EClinicalMedicine. 2020 Aug;25.

21. Central Statistical Office. 2010 Census of Population and Housing; Population and Demograhic projections 2011 - 2035. Zambia Cent Stat Off. 2013;142.

22. StataCorp. Stata Statistical Software: Release 14. College Station, TX: StataCorp LP. 2015;14.

23. World Health Organization. Director-General’s opening remarks at the media briefing on COVID-19 - 24 February 2020. 2020 Feb.

24. Furukawa NW, Furukawa NW, Brooks JT, Sobel J. Evidence Supporting Transmission of Severe Acute Respiratory Syndrome Coronavirus 2 while Presymptomatic or Asymptomatic. Emerg Infect Dis. 2020;26(7):E1–6.

25. World Health Organization. Global COVID-19 Clinical Platform Case Report Form (CRF) for Post COVID condition (Post COVID-19 CRF).. 2021 Feb.

26. Buonsenso D, Munblit D, De Rose C, Sinatti D, Ricchiuto A, Carfi A, et al. Preliminary Evidence on Long COVID in children. medRxiv. 2021;2021.01.23.21250375.

27. Sudre CH, Murray B, Varsavsky T, Graham MS, Penfold RS, Bowyer RC, et al. Attributes and predictors of long COVID. Nat Med. 2021 Mar;

28. Tenforde MW, Billig Rose E, Lindsell CJ, Shapiro NI, Files DC, Gibbs KW, et al. Characteristics of Adult Outpatients and Inpatients with COVID-19 — 11 Academic Medical Centers, United States, March–May 2020. MMWR Morb Mortal Wkly Rep. 2020;69(26):841–6.

29. Huang C, Huang L, Wang Y, Li X, Ren L, Gu X, et al. 6-month consequences of COVID-19 in patients discharged from hospital: a cohort study. Lancet. 2021 Jan;397(10270).

30. You J, Zhang L, Ni-jia-Ti M, Zhang J, Hu F, Chen L, et al. Anormal pulmonary function and residual CT abnormalities in rehabilitating COVID-19 patients after discharge. J Infect. 2020 Aug;81(2).

31. Zi-Wei Ye, Dong-Yan Jin. Diagnosis, treatment, control and prevention of SARS-CoV-2 and coronavirus disease 2019: back to the future. PMID. 2020 Apr;

32. Mahase E. Covid-19: Four in 10 people with evidence of past infection had no classic symptoms, study finds. BMJ. 2021 Feb;

